# COVID-19 vaccine effectiveness in children under 5 in the USA: a test-negative case-control study

**DOI:** 10.64898/2026.05.28.26354328

**Authors:** Rachel A. Silverman, Monica L. Ahrens, Meagan Helmick, Carla V. Finkielstein, Alasdair Cohen, Erica Short, Paige Bordwine

**Author notes:** **Address correspondence to:** Rachel Silverman, Center for Biostatistics and Health Data Science, Four Riverside Circle, Roanoke, VA 24016. [ ]. **Role of Funder/Sponsor:** The funders had no role in the design and conduct of this study.

## Abstract

**Background and Objectives:** SARS-CoV-2 (COVID-19) continues to mutate, circulate, and adversely impact health and quality of life. While COVID-19 vaccines remain safe and effective, uptake remains low, especially among children, the youngest of whom were not vaccine-eligible until after Omicron and are underrepresented in published research. This study estimated vaccine effectiveness (VE) among under-5-year-olds.

**Methods:** We used Virginia Department of Health surveillance data from June 2022 through October 2022 to conduct a test negative case-control study. We estimated VE derived from odds ratios (ORs) of reported infections using logistic regression among children aged 6-months to 5-years.

**Results:** Using the earliest positive (cases) or negative (controls) post-vaccine-eligible test results, the VE associated with two doses of a COVID-19 vaccine was 78% (95% CI=45%, 93%; p=0.004) in unadjusted analyses and 70% (95% CI=25%, 91%, p=0.023) when adjusting for age, sex, prior testing behavior, and prior reported infections. The adjusted VE was 74% (95% CI=28%, 94%; p=0.025) among those with no prior positives reported and 45% (95% CI=-302%, 97%; p=0.569) among those with a prior positive reported.

**Conclusions:** These results show that even though the vaccine was not closely matched to the dominant variants circulating during the time period analyzed, it was effective at reducing the risk of reported infections. This study adds to the body of knowledge on pediatric COVID-19 VE in an underrepresented age-group and in a rural region, illustrates the utility of surveillance data for evaluation, and can inform vaccine decisions to improve vaccine uptake for young children.

**What’s Known on This Subject:** SARS-CoV-2 (COVID-19) continues to mutate, circulate, and adversely impact health and quality of life. While research continues to show that COVID-19 vaccines remain safe and effective, uptake remains low, especially among young children who were not vaccine-eligible until after Omicron.

**What This Study Adds:** Children under-5 are underrepresented in published COVID-19 vaccine effectiveness research. This test negative case-control study estimates VE in this age-group following vaccine approval after the 2022 Omicron wave, using surveillance data from the Virginian Department of Health in Southwest Virginia.

## Background

While coronavirus disease 2019 (COVID-19) continues to adversely impact health and quality of life, ^1–4^ COVID-19 vaccines remain safe and effective at reducing illness and disease severity across populations. ^5–14^ However, SARS-CoV-2 continues to mutate, generating variants with variable transmissibility and disease severity, ^2, 15–18^ and it has been shown repeatedly that VE declines as immunity wanes and immune-escaping variants emerge, ^10, 11, 13, 14^ including among children. ^14^ Though at lower risk for serious illness than adults, some children do become severely ill^19–22^ and are at risk for post-acute sequelae of SARS-CoV-2 (or long-COVID). ^23–31^ Children are significant contributors to household transmission^32–35^ and illness can exacerbate health and economic disparities, ^36^ demonstrating the importance of reducing infection and illness risk across all age groups.

In early 2021, COVID-19 vaccines became available in the US to the general 16-and-older population, with lagging approval for pediatric age-groups (Figure 1). ^37–64^ It was not until after the initial Omicron wave during the 2021/2022 winter^41^, driving an explosion of viral diversity^40^ and leaving an estimated 75% of children with serologic evidence of a prior SARS-CoV-2 infection by February 2022, ^42^ that the vaccines for the youngest group (6 months to <5 years of age) were first approved on June 18^th^, 2022.

**Figure 1.**
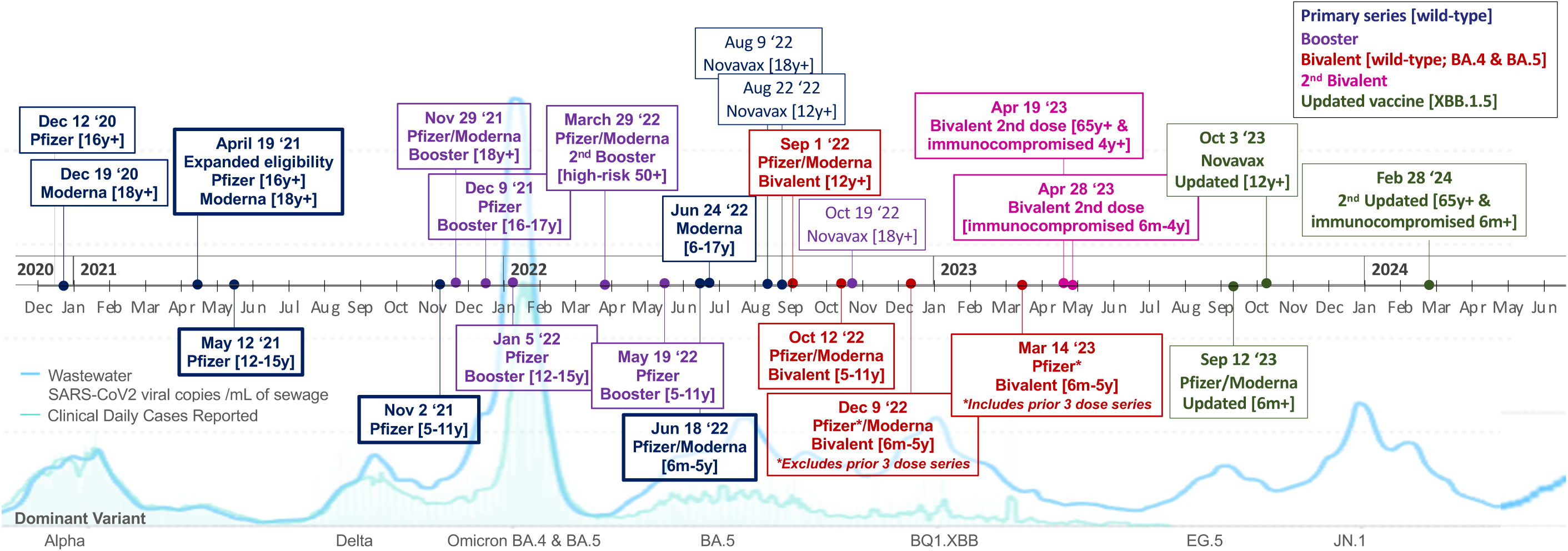
CDC COVID-19 vaccine recommendation timeline, superimposed over US reported daily cases (incidence) and SARS-CoV-2 wastewater concentrations (prevalence) from Biobot Analytics, Inc. (used with permission). Age-group and other eligibility criteria in brackets; vaccine type: primary series (bold border); 2020/2021-wild-type (blue), 2021/2022-wild-type-booster (purple), 2022/2023-wild-type/omicron bivalent-booster (red), second-bivalent-booster (pink), and 2023/2024-monovalent XBB.1.5 (green). **Alt Text:** Timeline of COVID-19 Pfizer-BioNTech (Pfizer), Moderna, and Novavax CDC recommendations by age-group and temporal trends in SARS-CoV-2 incidence and prevalence from clinical case reports and waste-water surveillance. Note that the CDC approved the initial primary series for children 6 months to 5 years in June 2022, after the Omicron wave during the prior winter, lagging over a year behind adult and older adolescent approval in April and May, 2021, and 7 months after the 5-11 year old age-group approval in November, 2021.

COVID-19 Vaccines for SARS-CoV-2 are recommended and approved for everyone over 6 months of age by the FDA, CDC, ACIP, and the AAP, ^47, 48, 51, 52, 60, 61, 65^ including with vaccines released annually in the fall that are updated to target recently dominating variants. ^38, 39, 64, 66^ However, vaccine uptake among pediatric populations remains extremely low, particularly in marginalized communities and rural areas. ^67–70^ Vaccine hesitancy among parents has increased, especially for COVID-19. ^71^ According to the CDC, approximately 14.9% of children 6-months to 17-years old were up-to-date with COVID-19 vaccines by the end of the 2023-2024 season, declining to 13.2% for 2024-2025. ^70^ Coverage was disproportionately low among rural pediatric populations, at 8.4% and 7.5% by end of these seasons, respectively. ^70^ Among children 6 months to 4 years, coverage was much lower at 6.7% and 5.9%, respectively. ^70^

While there is substantial research on the VE for the 5 and up age-groups, ^6–8, 72–94^ only a few published studies exist that focus on children under 5 years old, ^95–100^ and fewer focus on VE against reported infections^96, 97^ vs. hospitalizations for higher risk individuals. ^98, 100^ Evaluating VE in diverse contexts informs policy makers, healthcare providers, and families in making vaccination decisions. Here, we estimated VE against reported COVID-19 infection among vaccine-eligible children under 5 using VDH surveillance data from rural Southwest Virginia.

## Methods

We conducted a test-negative case-control study to estimate VE of the COVID-19 mRNA primary series for children aged 6 months to under 5 years between June 18 and October 31, 2022 — the date after which negative tests were no longer reported to VDH. Data were restricted to Southwest Virginia (SWVA) to ensure consistency and focus on a rural population.

This study was approved by the Institutional Review Boards (IRBs) of Virginia Tech (IRB#: 20-852) and the Virginia Department of Health (IRB#: 70046). The study was determined to be public health surveillance as defined in the US Department of Health and Human Services, Title 45 Code of Federal Regulations, §46 Protection of Human Subjects (45 CFR §46.102[l]), and thus informed consent was not required.

### Data sources and linkage

All data were routinely collected and provided by VDH. COVID-19 case and laboratory data are stored in the Virginia Electronic Disease Surveillance System (VEDSS); vaccination data are stored in the Virginia Immunization Information System (VIIS). VDH linked these datasets via 85% confidence fuzzy matching on first name, last name, and date of birth, restricting records to individuals aged 6 months to 18 years with at least one test before October 31, 2022, living in SWVA. A unique identifier was created prior to de-identification to allow validation while protecting confidentiality.

### Selection Criteria

Selection of records retained for analysis is outlined in Figure 2. Selected records included children aged 6 months to under 5 years with samples collected after their vaccine eligibility date, defined as June 22, 2022 (the earliest vaccine received in this cohort), or the date they turned 6 months old, up to October 31, 2022. Five controls were excluded for residing outside SWVA, and 3 individuals (1 case, 2 controls) were excluded due to invalid merged vaccine information in which their names and dates of births matched, but their age did not (were 10-14 years old), with a vaccine type inconsistent with their age and an administered date prior to eligibility.

**Figure 2.**
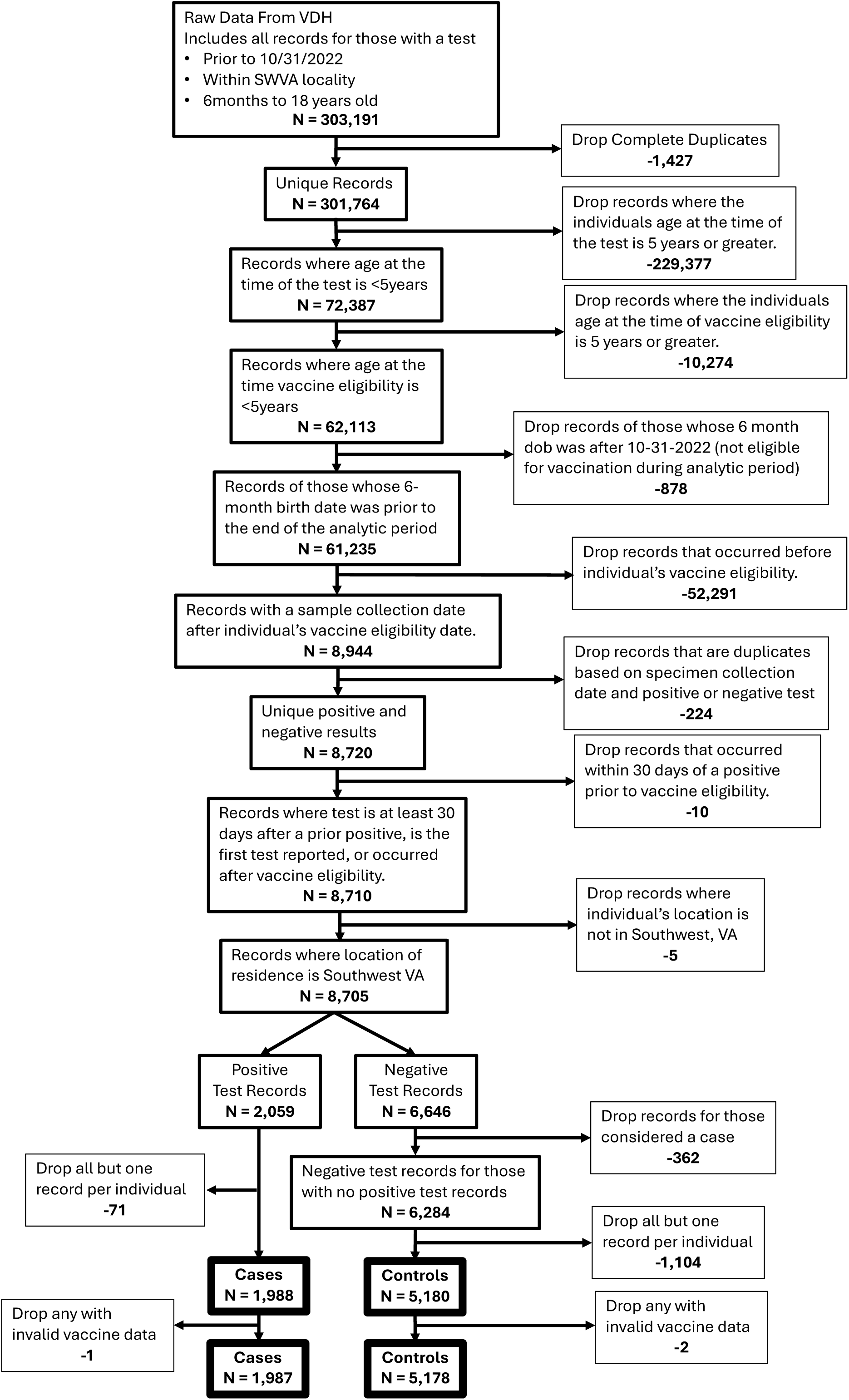
Inclusion and exclusion criteria flow-chart for selection into test-negative case-control cohort. **Alt Text:** Flow chart for selection into analytic case-control study cohort. From 303,191 records, excluding duplicates and those that do not meet inclusion criteria. Ultimately identifying 1,987 cases and 5,178 controls included in the analysis cohort.

### Case and control definitions

Cases were defined as those who had at least one positive test reported after they were eligible for vaccination. Controls were defined as those who had at least one negative test (and no positive tests) reported after vaccination eligibility. Cases and Controls were selected irrespective of the number and results of tests dated prior to vaccine eligibility.

### Variable Definitions

Age was calculated from date of birth to sample collection date and defined as continuous (days) and categorical by year of age (<1, 1 to <2, 2 to <3, 3 to 4, 4 to <5 years). Sex was binary (male vs. female). Race and ethnicity were combined into one categorical variable: Hispanic (irrespective of race), and non-Hispanic White, Black, Asian, or other; the “other” category included American Indian or Alaska Native and Native Hawaiian or Pacific Islander due to small numbers. Individuals reporting multiple races were assigned to the group with the smallest count in the dataset^101^. Health district (Alleghany, Central Virginia, Cumberland Plateau, Lenowisco, Mount Rogers, New River, Pittsylvania, Roanoke City, and West Piedmont) ^102^ was assigned based on residence at the time of the test; for missing counties, assignment was based on ZIP code, with cross-county ZIPs assigned to the most populous county.

Prior reported infections were positive results reported to VDH at least 30 days apart and before vaccine eligibility; when multiple positives occurred within 30 days, only the earliest was retained. Prior test-seeking behavior was categorized as: 1) first-ever test (no tests prior to eligibility), 2) sporadic (1–3 tests), or 3) frequent (≥4 tests), excluding tests within 30 days of a prior positive. In multivariable regression, behavior was defined by the number of prior negative tests.

Vaccine status — the primary predictor — was categorized by number of doses (1–3), vaccine type (Pfizer-BioNTech or Moderna), and time since last dose. Because many controls had multiple negative tests, using vaccine status at the earliest test would underestimate VE (less opportunity to vaccinate), while using the most-recent test would overestimate it (more opportunity). To bound the VE estimates:

- For cases, the vaccine status at their first positive test post-vaccine eligibility was used in all analyses.
- For controls, separate analyses were conducted in which the vaccine status at the 1) earliest and 2) most-recent negative test post-vaccine eligibility was used.

To allow time for immunity induction, doses administered within 7 days of the test were grouped with the previous dose category (e.g., first dose within 7 days = unvaccinated; second dose within 7 days = 1 dose; third dose within 7 days = 2 doses).

### Statistical Analyses

Descriptive statistics (mean, min, max, median for continuous; number and percent for categorical) were calculated for the full cohort and by case/control status at the earliest eligible test. Vaccine type and dose counts are described for cases and controls under both selection scenarios (earliest and most-recent).

Unadjusted and adjusted odds ratios (OR) comparing vaccinated (by dose count) to unvaccinated were estimated via logistic regression (α = 0.05), adjusting for age, sex, race/ethnicity, prior positive infections, time since prior positives, and prior test-seeking behavior. AIC determined the optimal age parameterization; Health District was included in sensitivity analyses. Vaccine types were combined due to small numbers. Time since last dose was included to assess waning immunity. VE was calculated as [VE = (1 − OR) × 100].

All data cleaning and analyses were conducted using RStudio. ^103, 104^ Some R code was partially generated using ChatGPT^105^ and all code was manually reviewed to confirm accuracy. R packages include tidyverse, ^106^ gtsummary, ^107^ and epikit. ^108^ The complete R scripts and a limited dataset are available in the supplemental material.

## Results

### Demographics

The study included 7,165 individuals (5,178 controls and 1,987 cases; Table 1). The overall mean age was 2.6 years; cases were slightly younger than controls (2.32 vs. 2.68 years) and had nearly twice the proportion of infants under 1 year (23% vs. 13%). Sex distribution was similar between groups (47% female overall, <1% missing). Race/ethnicity data were missing in 44% of controls vs. 9.8% of cases — a disparity arising because VDH routinely collected this information during case investigations for positive tests but not for negative ones. Among those with data, 3,924 (83.6%) identified as White and 500 (10.7%) as Black/African American, slightly lower than the 14.1% census estimate of the proportion of children under 5 who are non-Hispanic Black/African American in Southwest Virginia in 2022. ^109^ Given the large differential missingness, race/ethnicity was excluded from regression analyses.

**Table 1.** Characteristics of study cohort by overall and by case and control* status. **Alt Text:** Descriptive statistics of study cohort characteristics, including age, sex, race/ethnicity, prior positive, negative, and any tests reported. Differences were observed with controls having greater proportion of individuals with prior tests (positive and negative), missing more race/ethnicity data, and having a smaller proportion of children under 1 years-old than cases.

| **Characteristic** |  | **Overall (N=7,165)** | **Cases (N=1,987)** | **Controls (N=5,178)** |
| --- | --- | --- | --- | --- |
| **Age at test (years)** | Mean (SD) | 2.58 (1.36) | 2.32 (1.38) | 2.68 (1.33) |
|  | Median [Min, Max] | 2.52 [0.5, 5] | 2.04 [0.5, 5] | 2.69 [0.5, 5] |
| **Age (years) Groups** | 6m to <1yr | 1,127 (15.7%) | 467 (23.5%) | 660 (12.8%) |
|  | 1 yrs | 1,757 (24.5%) | 505 (25.4%) | 1,252 (24.2%) |
|  | 2 yrs | 1,357 (18.9%) | 343 (17.3%) | 1,014 (19.6%) |
|  | 3 yrs | 1,466 (20.5%) | 335 (16.9%) | 1,131 (21.8%) |
|  | 4 yrs | 1,458 (20.3%) | 337 (17.0%) | 1,121 (21.6%) |
| **Sex** | Female | 3,390 (47.3%) | 932 (46.9%) | 2,458 (47.5%) |
|  | Male | 3,734 (52.1%) | 1,053 (53.0%) | 2,681 (51.8%) |
|  | Unknown/missing | 41 (0.6%) | 2 (0.1%) | 39 (0.8%) |
| **Race/Ethnicity\*\*** | White | 3,924 (54.7%; 83.6%) | 1,414 (71.2%; 78.9%) | 2,510 (48.4%; 86.5%) |
|  | Black | 500 (7.0%; 10.7%) | 237 (11.9%; 13.2%) | 263 (5.1%; 9.1%) |
|  | Asian | 43 (0.6%; 0.9%) | 20 (1.0%; 1.1%) | 23 (0.4%; 0.8%) |
|  | Hispanic | 96 (1.3%; 2.0%) | 96 (4.8%; 5.4%) | 0 (0%; 0%) |
|  | Other | 128 (1.8%; 2.7%) | 26 (1.3%; 1.5%) | 102 (2.0%; 3.5%) |
|  | Unknown/missing | 2,474 (34.5%; na) | 194 (9.8%; na) | 2,280 (44.0%; na) |
| **Prior Positive Test Reported** | none | 5,959 (83.2%) | 1,742 (87.7%) | 4,217 (81.4%) |
|  | 1 or more | 1,206 (16.8%) | 245 (12.3%) | 961 (18.6%) |
| **Prior Tests Reported (any)** | None | 3,227 (45.0%) | 1,082 (54.5%) | 2,145 (41.4%) |
|  | 1 to 3 | 3,347 (46.7%) | 779 (39.2%) | 2,568 (49.6%) |
|  | 4 or more | 591 (8.2%) | 126 (6.3%) | 465 (9.0%) |
| **Prior Negative Test Reported** | None | 3,654 (51.0%) | 1,174 (59.1%) | 2,480 (47.9%) |
|  | 1 to 3 | 3,092 (43.2%) | 721 (36.3%) | 2,371 (45.8%) |
|  | 4 or more | 419 (5.8%) | 92 (4.6%) | 327 (6.3%) |
| *As of the date of their earliest post-vaccine eligible test (for controls) |  |  |  |  |
| **N (% of total including unknown; % of total excluding unknown) |  |  |  |  |

### Prior testing and positives

Overall, 17% of the cohort had at least one prior infection reported before vaccine eligibility, with a greater proportion among controls than cases (18% vs. 12%; Table 1). Forty-five percent had no prior tests reported, with fewer controls than cases in this category (41% vs. 54%). In summary, controls were more frequently tested and more likely to have had a prior positive than cases.

### Vaccine coverage

Of the controls, 839 (16%) had more than one post-eligible negative test; differences between earliest and most-recent control scenarios are driven by these individuals. Overall vaccine coverage was very low (<2%; Table 2). Of cases, 15 (0.9%) had received at least one dose before their positive test, compared to 92 (1.8%) and 99 (1.9%) of controls at earliest and most-recent tests, respectively. After applying the 7-day exclusion window, 8 (0.4%) cases had received 1 dose and 4 (0.2%) had received 2 doses; controls had 39 (0.8%) and 37 (0.7%) with 1 dose and 46 (0.9%) and 55 (1.1%) with ≥2 doses (earliest and most-recent, respectively). Of the 12 analytically vaccinated cases, 3 (0.2%) received Moderna and 9 (0.3%) received Pfizer-BioNTech. Of the controls, as of the earliest test, 29 (0.6%) received Moderna and 56 (1.1%) received Pfizer-BioNTech; as of the most-recent test, 32 (0.6%) received Moderna and 60 (1.1%) received Pfizer-BioNTech at least 7 days prior to their test.

**Table 2.**
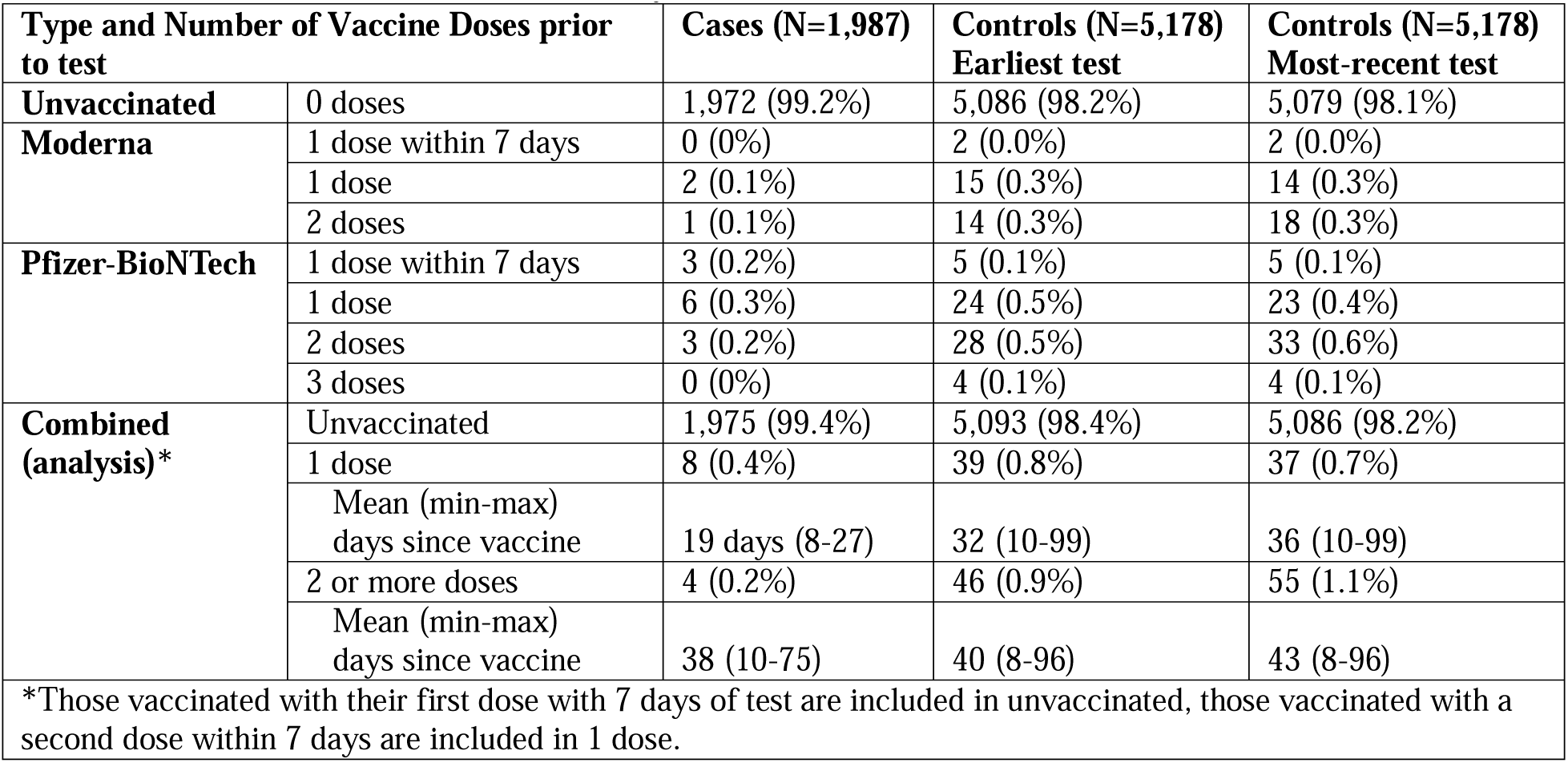
Vaccine status* at the time of the analytic test for cases and controls. **Alt Text:** Vaccine status frequencies at the time of the analytic test for cases and controls, by type (Moderna vs. Pfizer BioNTech), number of doses, and mean number of days since the most recent vaccine.

| **Type and Number of Vaccine Doses prior to test** |  | **Cases (N=1,987)** | **Controls (N=5,178) Earliest test** | **Controls (N=5,178) Most-recent test** |
| --- | --- | --- | --- | --- |
| **Unvaccinated** | 0 doses | 1,972 (99.2%) | 5,086 (98.2%) | 5,079 (98.1%) |
| **Moderna** | 1 dose within 7 days | 0 (0%) | 2 (0.0%) | 2 (0.0%) |
|  | 1 dose | 2 (0.1%) | 15 (0.3%) | 14 (0.3%) |
|  | 2 doses | 1 (0.1%) | 14 (0.3%) | 18 (0.3%) |
| **Pfizer-BioNTech** | 1 dose within 7 days | 3 (0.2%) | 5 (0.1%) | 5 (0.1%) |
|  | 1 dose | 6 (0.3%) | 24 (0.5%) | 23 (0.4%) |
|  | 2 doses | 3 (0.2%) | 28 (0.5%) | 33 (0.6%) |
|  | 3 doses | 0 (0%) | 4 (0.1%) | 4 (0.1%) |
| **Combined (analysis)\*** | Unvaccinated | 1,975 (99.4%) | 5,093 (98.4%) | 5,086 (98.2%) |
|  | 1 dose | 8 (0.4%) | 39 (0.8%) | 37 (0.7%) |
|  | Mean (min-max) days since vaccine | 19 days (8-27) | 32 (10-99) | 36 (10-99) |
|  | 2 or more doses | 4 (0.2%) | 46 (0.9%) | 55 (1.1%) |
|  | Mean (min-max) days since vaccine | 38 (10-75) | 40 (8-96) | 43 (8-96) |
| *Those vaccinated with their first dose with 7 days of test are included in unvaccinated, those vaccinated with a second dose within 7 days are included in 1 dose. |  |  |  |  |

Due to the small number of vaccinated individuals, vaccine types (Pfizer-BioNTech and Moderna) were combined, and vaccine status was defined by the number of doses received at least 7 days prior to the test (see Table 2). Mean days since vaccination for cases were 19 (1 dose) and 38 (2 doses); for controls, 32 and 40 days (earliest test) and 36 and 43 days (most-recent test) for 1 and ≥2 doses, respectively.

### Vaccine Effectiveness and Logistic Regression Results

Unadjusted and adjusted VE results are presented in Table 3. One dose was associated with an unadjusted VE of 47% (95% CI: −7%, 77%; p=0.100) using the earliest test and 44% (−14%, 76%; p=0.130) using the most-recent test. Two or more doses yielded an unadjusted VE of 78% (95% CI: 45%, 93%; p=0.004) and 81% (54%, 94%; p=0.001), respectively. After adjustment for age, sex, prior positivity, and prior testing frequency, the estimates were modestly attenuated: 1 dose: 32% (−38%, 71%; p=0.316) and 29% (−46%, 70%; p=0.382); ≥2 doses: 70% (26%, 91%; p=0.021) and 76% (40%, 93%; p=0.007), for earliest and most-recent controls, respectively.

**Table 3.**
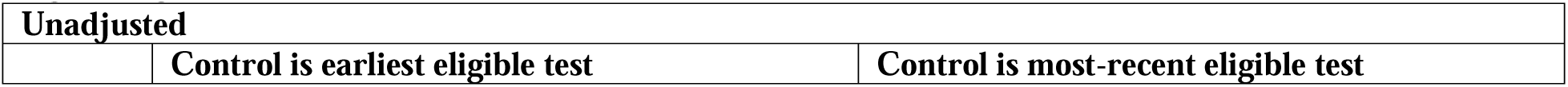
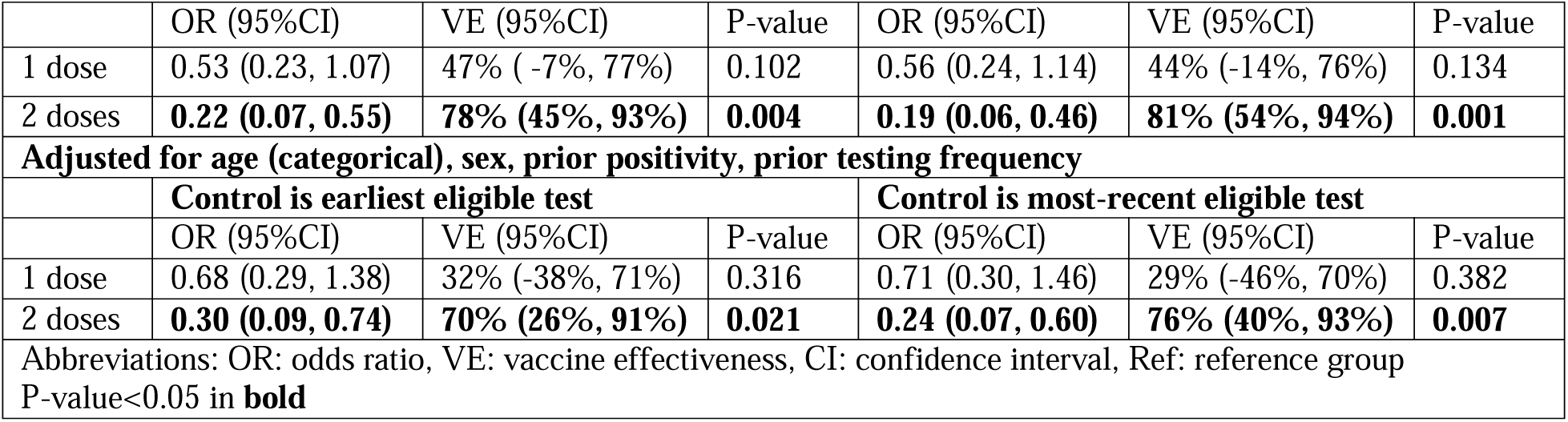
Vaccine effectiveness* by number of doses calculated from odds ratios from unadjusted and adjusted logistic regression. **Alt Text:** Vaccine effectiveness estimates by number of doses calculated from odds ratios from unadjusted and adjusted logistic regression. Adjusted analyses found the VE was attenuated relative to unadjusted results but remained statistically significantly effective when comparing those with 2 doses to unvaccinated.

Categorical age was preferred based on AIC; results were robust to alternative age parameterizations (supplement Table S1). Full multivariable results are in Table 4. Increasing age, prior negative tests, and prior positives were each significantly associated with lower odds of a reported positive test (p<0.05); sex was not. Results were consistent across control selection scenarios and age parameterizations (supplement Table S1).

**Table 4.**
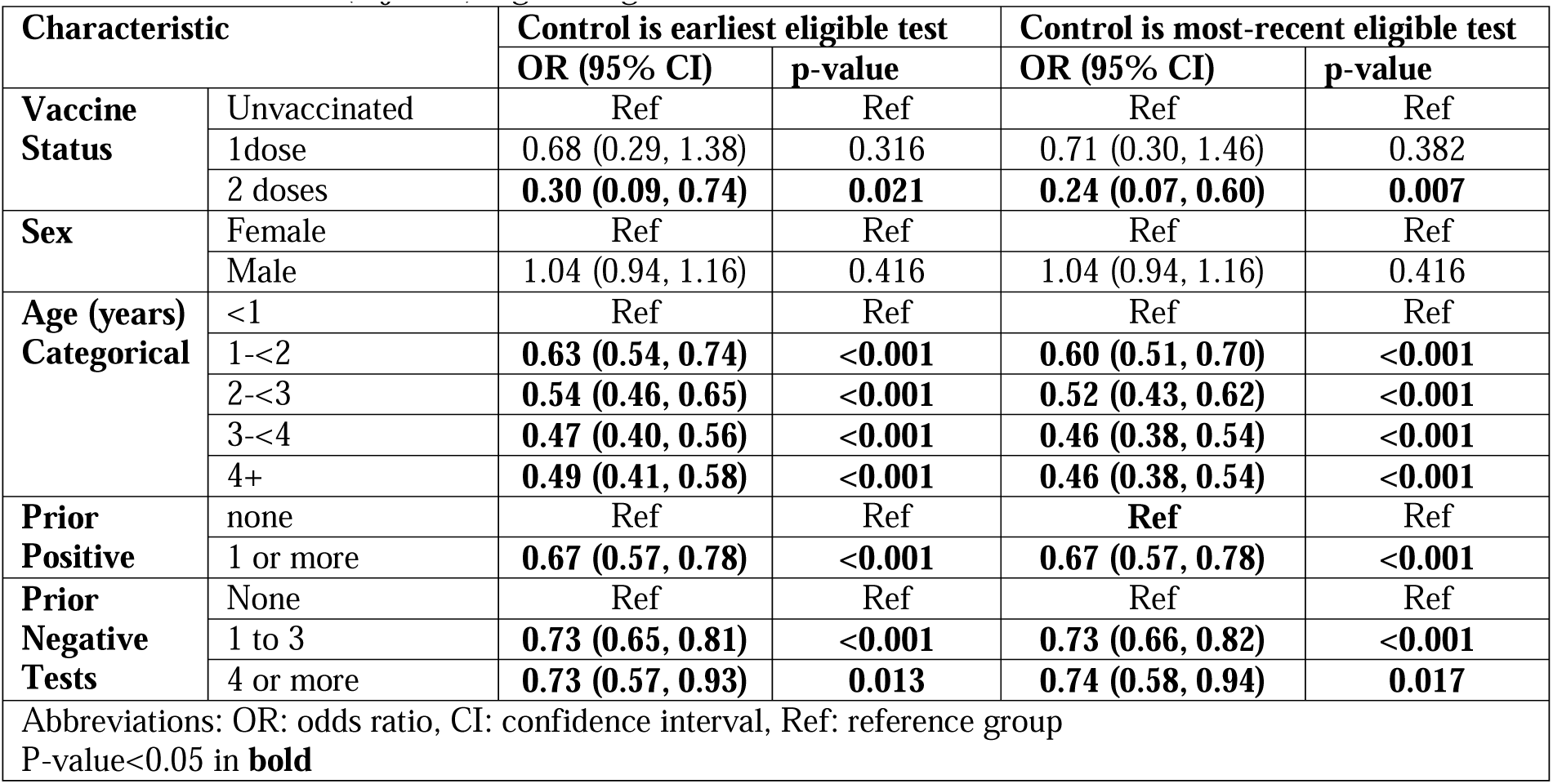
Multivariable (adjusted) logistic regression model results. **Alt Text:** Multivariable (adjusted) logistic regression model results, presenting odds ratios for a positive reported test for all variables included in the model: vaccine status, sex, age, prior positive test, and prior negative tests.

Stratified analyses (Table 5) showed that among children with no prior positive tests, 2 doses were associated with an OR of 0.26 (95% CI: 0.06, 0.72; p=0.025), corresponding to a VE of 74% (95% CI: 28%, 94%). Among those with a prior positive, the OR associated with 2 doses was 0.55 (95% CI: 0.03, 3.02; p=0.569), yielding a non-significant VE of 45% (95% CI: −302%, 97%). Younger age was associated with higher odds of infection in both strata. Prior negative tests were associated with lower odds only among those without a prior positive. Results were consistent across control scenarios and age parameterizations (supplement Tables S2, S3). The vaccine status effect was similar when health district was added to the multivariate models (see supplement: table S4).

**Table 5.**
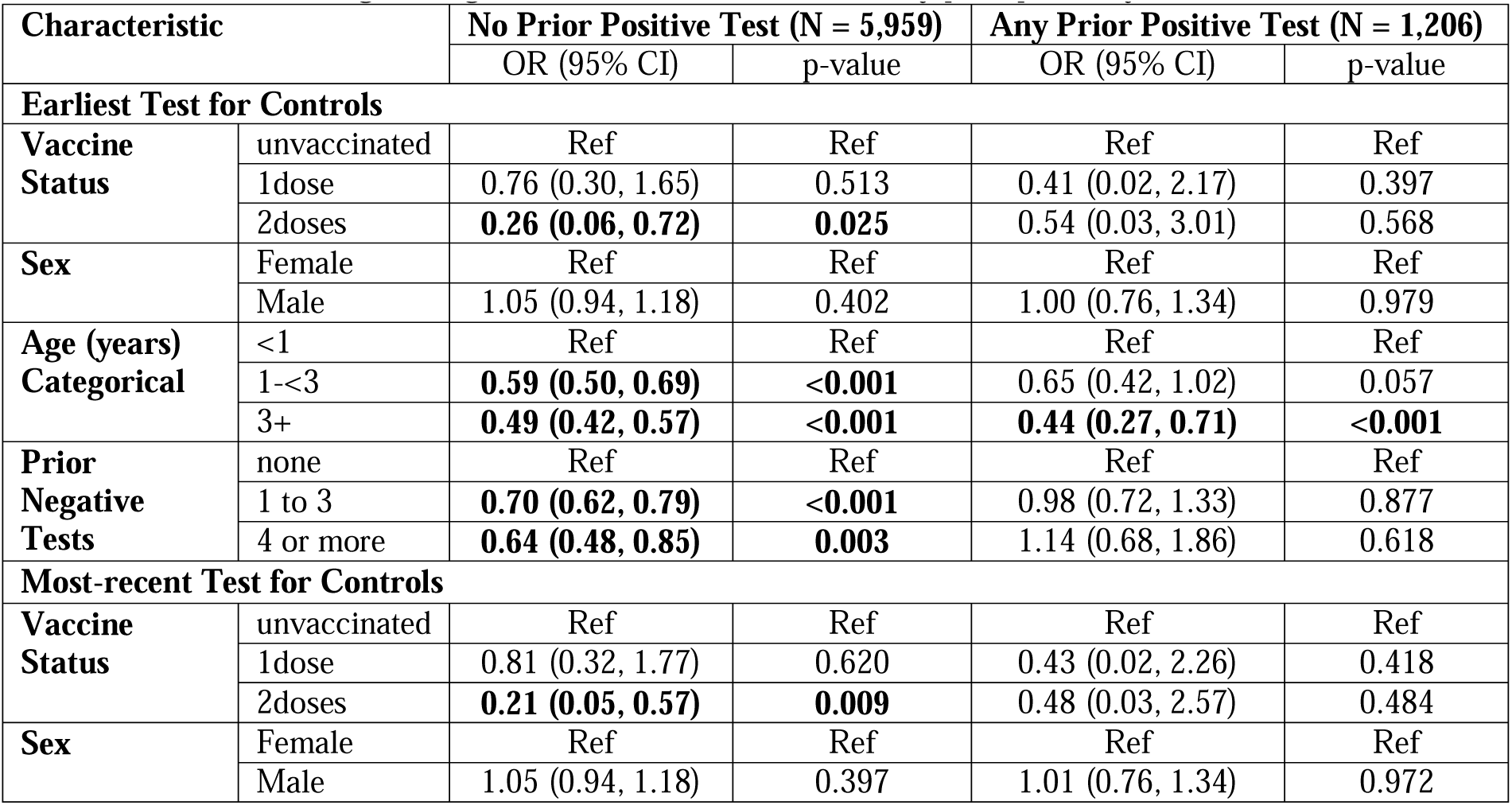
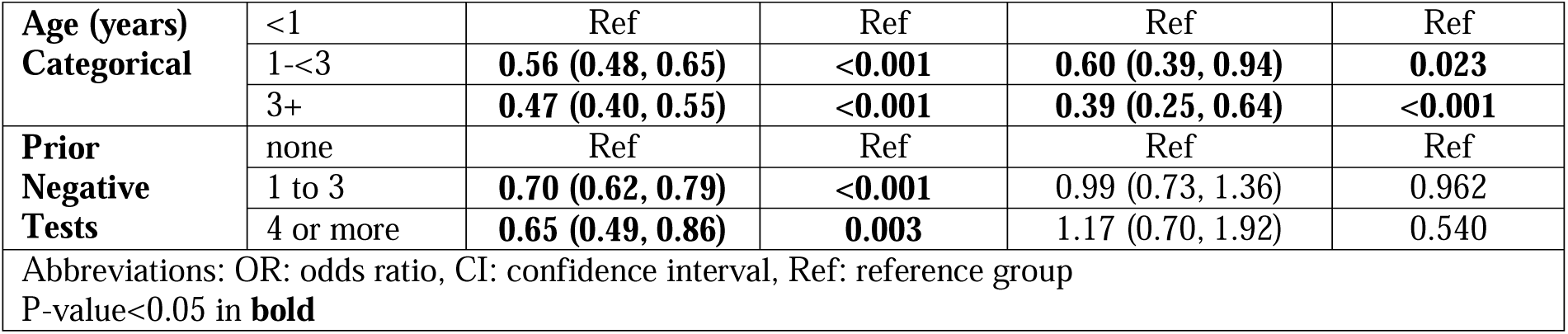
Multivariable logistic regression model results, stratified by prior positivity. **Alt Text:** Multivariable (adjusted) logistic regression model results stratified by prior positive test, presenting odds ratios for a positive reported test for all variables included in the model: vaccine status, sex, age, and prior negative tests. Note that the vaccine effectiveness was much greater among those with no prior positive test compared to those with a prior positive test.

## Discussion

This study demonstrated that among children under 5 who became vaccine-eligible after the Omicron wave, vaccination — particularly ≥2 doses — significantly reduced reported infection risk. Prior infection was itself protective and attenuated the observed VE, consistent with the literature that natural infection confers partial immunity to reinfection^110^. Higher pre-eligibility testing frequency was also associated with lower odds of a subsequent reported infection, but only among those without a prior positive — suggesting a systematic difference in risk between frequent and infrequent testers in this subgroup.

This study has several limitations. First, case and control classification relies solely on tests reported to VDH; rapid test results and unreported infections are not captured, and the test-negative design overrepresents symptomatic illness while underrepresenting mild or asymptomatic infections. Second, waning immunity was not modeled, though this is likely negligible given the brief analytic window and limited variant diversity during this period (Figure 1). Third, race and ethnicity could not be included in regression due to high missingness among controls. Fourth, low vaccine uptake precluded type-specific VE analyses. Fifth, VE against severity gradients or long-COVID could not be assessed. Sixth, post-eligibility test-seeking behavior — which may differ between vaccinated and unvaccinated — was not accounted for. Finally, because ORs overestimate relative risk when disease prevalence is high, VE estimates are likely conservative. ^111^

## Conclusions

This study found that vaccination was effective at reducing medically attended COVID-19 infections in this cohort. By focusing on the youngest children in an underrepresented rural region with disproportionately low uptake, this study contributes meaningfully to the pediatric COVID-19 VE literature and demonstrates the value of positive and negative surveillance data for real-world vaccine evaluation. While novel variants continue to emerge, ^112^ and children continue to have low vaccine uptake for COVID-19 vaccines^70^ despite the recommendations by the CDC^66^ and AAP, ^113^ these results can help inform vaccine decisions and improve vaccine uptake for young children.

## Data Availability

All data produced in the present study are available upon reasonable request to the authors

## Abbreviations

AAP: American Academy of Pediatrics
ACIP: Advisory Committee on Immunization Practices
AIC: Akaike information criterion
CDC: Centers for Disease Control and Prevention
CI: Confidence interval
COVID-19: Coronavirus disease 2019
FDA: Food and Drug Administration
IRB: Institutional Review Boards
OR: Odds ratio
SARS-CoV-2: Severe acute respiratory syndrome coronavirus 2
SWVA: Southwest Virginia
VDH: Virginia Department of Health
VE: Vaccine effectiveness
VEDSS: Virginia Electronic Disease Surveillance System
VIIS: Virginia Immunization Information System

## Contributors Statement Page

Dr. Rachel Silverman conceptualized and designed the study, conducted all data analyses, and drafted and finalized the manuscript.

Dr. Monica Ahrens consulted on and validated the methods and critically reviewed and revised the manuscript.

Dr. Alasdair Cohen consulted on data analysis methods and critically reviewed and revised the manuscript.

Dr. Carla Finkielstein validated the data tables and critically reviewed and revised the manuscript.

Dr. Meagan Helmick was involved in data collection and validation, consulted on the methods and critically reviewed and revised the manuscript.

Erica Short and Paige Bordwine were involved in data collection and validation and critically reviewed and revised the manuscript.

All authors approved the final manuscript as submitted and agree to be accountable for all aspects of the work.

## Acknowledgements

Thank you to the Virginia Department of Health and the Center for Biostatistics and Health Data Science art Virginia Tech.

