## Supplement for "COVID-19 vaccine effectiveness in children under 5 in the USA: a test-negative case-control study"

**Table S1.** Multivariable logistic regression results including age as continuous (model 1), in 5 categories (model 2), and as 3 categories (model 3).

| **Model 1** | | **Control is earliest eligible test** | | **Control is most-recent eligible test** | |
| --- | --- | --- | --- | --- | --- |
| **Characteristic** | | **OR (95% CI)** | **p-value** | **OR (95% CI)** | **p-value** |
| Vaccine  Status | Unvaccinated | Ref | Ref | Ref | Ref |
|  | 1dose | 0.65 (0.28, 1.33) | 0.274 | 0.68 (0.29, 1.40) | 0.331 |
|  | 2 doses | **0.30 (0.09, 0.75)** | **0.023** | **0.24 (0.07, 0.60)** | **0.007** |
| Sex | Female | Ref | Ref | Ref | Ref |
|  | Male | 1.05 (0.94, 1.16) | 0.403 | 1.05 (0.94, 1.16) | 0.412 |
| Age (years)  Continuous | One unit increase | **0.84 (0.81, 0.88)** | **<0.001** | **0.84 (0.80, 0.87)** | **<0.001** |
| Prior Positive | none | Ref | Ref | Ref | Ref |
|  | 1 or more | **0.66 (0.56, 0.76)** | **<0.001** | **0.66 (0.56, 0.76)** | **<0.001** |
| Prior Negative Tests | none | Ref | Ref | Ref | Ref |
|  | 1 to 3 | **0.71 (0.63, 0.79)** | **<0.001** | **0.71 (0.64, 0.79)** | **<0.001** |
|  | 4 or more | **0.70 (0.54, 0.89)** | **0.004** | **0.71 (0.55, 0.90)** | **0.006** |
| **Model 2** | | **Control is earliest eligible test** | | **Control is most-recent eligible test** | |
| **Characteristic** | | **OR (95% CI)** | **p-value** | **OR (95% CI)** | **p-value** |
| Vaccine  Status | Unvaccinated | Ref | Ref | Ref | Ref |
|  | 1dose | 0.68 (0.29, 1.38) | 0.316 | 0.71 (0.30, 1.46) | 0.382 |
|  | 2 doses | **0.30 (0.09, 0.74)** | **0.021** | **0.24 (0.07, 0.60)** | **0.007** |
| Sex | Female | Ref | Ref | Ref | Ref |
|  | Male | 1.04 (0.94, 1.16) | 0.416 | 1.04 (0.94, 1.16) | 0.416 |
| Age (years)  Categorical | <1 | Ref | Ref | Ref | Ref |
|  | 1-<2 | **0.63 (0.54, 0.74)** | **<0.001** | **0.60 (0.51, 0.70)** | **<0.001** |
|  | 2-<3 | **0.54 (0.46, 0.65)** | **<0.001** | **0.52 (0.43, 0.62)** | **<0.001** |
|  | 3-<4 | **0.47 (0.40, 0.56)** | **<0.001** | **0.46 (0.38, 0.54)** | **<0.001** |
|  | 4+ | **0.49 (0.41, 0.58)** | **<0.001** | **0.46 (0.38, 0.54)** | **<0.001** |
| Prior Positive | none | Ref | Ref | Ref | Ref |
|  | 1 or more | **0.67 (0.57, 0.78)** | **<0.001** | **0.67 (0.57, 0.78)** | **<0.001** |
| Prior Negative Tests | None | Ref | Ref | Ref | Ref |
|  | 1 to 3 | **0.73 (0.65, 0.81)** | **<0.001** | **0.73 (0.66, 0.82)** | **<0.001** |
|  | 4 or more | **0.73 (0.57, 0.93)** | **0.013** | **0.74 (0.58, 0.94)** | **0.017** |
| **Model 3** | | **Control is earliest eligible test** | | **Control is most-recent eligible test** | |
| **Characteristic** | | **OR (95% CI)** | **p-value** | **OR (95% CI)** | **p-value** |
| Vaccine  Status | Unvaccinated | Ref | Ref | Ref | Ref |
|  | 1dose | 0.67 (0.29, 1.38) | 0.315 | 0.71 (0.31, 1.46) | 0.389 |
|  | 2 doses | **0.30 (0.09, 0.74)** | **0.021** | **0.24 (0.07, 0.60)** | **0.007** |
| Sex | Female | Ref | Ref | Ref | Ref |
|  | Male | 1.04 (0.94, 1.16) | 0.428 | 1.04 (0.94, 1.16) | 0.425 |
| Age (years)  Categorical | <1 | Ref | Ref | Ref | Ref |
|  | 1-<3 | **0.59 (0.51, 0.69)** | **<0.001** | **0.56 (0.49, 0.65)** | **<0.001** |
|  | 3+ | **0.48 (0.41, 0.56)** | **<0.001** | **0.46 (0.39, 0.53)** | **<0.001** |
| Prior Positive | none | Ref | Ref | Ref | Ref |
|  | 1 or more | **0.67 (0.57, 0.78)** | **<0.001** | **0.67 (0.58, 0.79)** | **<0.001** |
| Prior Negative Tests | None | Ref | Ref | Ref | Ref |
|  | 1 to 3 | **0.73 (0.65, 0.81)** | **<0.001** | **0.73 (0.65, 0.82)** | **<0.001** |
|  | 4 or more | **0.72 (0.56, 0.92)** | **0.009** | **0.73 (0.57, 0.93)** | **0.012** |
| Abbreviations: OR: odds ratio, CI: confidence interval, Ref: reference group  P-value<0.05 in **bold** | | | | | |

**Table S2.** Multivariable logistic regression model results, stratified by prior positivity, including age as continuous (model 1), in 5 categories (model 2), and as 3 categories (model 3), using earliest post-vaccine eligible negative for controls.

| **Characteristic** | | **No Prior Positive Test (N = 5,959)** | | **Any Prior Positive Test (N = 1,206)** | |
| --- | --- | --- | --- | --- | --- |
| **Model 1** | | **OR (95% CI)** | **p-value** | **OR (95% CI)** | **p-value** |
| Vaccine Status | unvaccinated | Ref | Ref | Ref | Ref |
|  | 1dose | 0.74 (0.30, 1.62) | 0.483 | 0.38 (0.02, 2.02) | 0.358 |
|  | 2doses | **0.26 (0.06, 0.74)** | **0.027** | 0.56 (0.03, 3.09) | 0.582 |
| Sex | Female | Ref | Ref | Ref | Ref |
|  | Male | 1.05 (0.94, 1.18) | 0.370 | 1.00 (0.76, 1.33) | 0.975 |
| Age (years)  Continuous | One unit increase | **0.85 (0.81, 0.89)** | **<0.001** | **0.80 (0.71, 0.90)** | **<0.001** |
| Prior Negative Tests | none | Ref | Ref | n/a | n/a |
|  | 1 to 3 | **0.67 (0.60, 0.76)** | **<0.001** | 0.99 (0.72, 1.35) | 0.927 |
|  | 4 or more | **0.62 (0.46, 0.82)** | **0.001** | 1.15 (0.69, 1.89) | 0.578 |
| **Model 2** | | **OR (95% CI)** | **p-value** | **OR (95% CI)** | **p-value** |
| Vaccine Status | unvaccinated | Ref | Ref | Ref | Ref |
|  | 1dose | 0.77 (0.31, 1.67) | 0.534 | 0.40 (0.02, 2.12) | 0.382 |
|  | 2doses | **0.26 (0.06, 0.72)** | **0.025** | 0.57 (0.03, 3.19) | 0.603 |
| Sex | Female | Ref | Ref | Ref | Ref |
|  | Male | 1.05 (0.94, 1.18) | 0.400 | 1.00 (0.76, 1.33) | 0.982 |
| Age (years) Categorical | <1 | Ref | Ref | Ref | Ref |
|  | 1-<2 | **0.62 (0.52, 0.74)** | **<0.001** | 0.72 (0.46, 1.15) | 0.169 |
|  | 2-<3 | **0.54 (0.45, 0.65)** | **<0.001** | **0.55 (0.33, 0.92)** | **0.022** |
|  | 3-<4 | **0.47 (0.39, 0.56)** | **<0.001** | **0.49 (0.29, 0.83)** | **0.007** |
|  | 4+ | **0.50 (0.42, 0.61)** | **<0.001** | **0.38 (0.22, 0.65)** | **<0.001** |
| Prior Negative Tests | none | Ref | Ref | Ref | Ref |
|  | 1 to 3 | **0.70 (0.62, 0.79)** | **<0.001** | 0.99 (0.73, 1.36) | 0.959 |
|  | 4 or more | **0.65 (0.49, 0.87)** | **0.004** | 1.18 (0.70, 1.93) | 0.530 |
| **Model 3** | | **OR (95% CI)** | **p-value** | **OR (95% CI)** | **p-value** |
| Vaccine Status | unvaccinated | Ref | Ref | Ref | Ref |
|  | 1dose | 0.76 (0.30, 1.65) | 0.513 | 0.41 (0.02, 2.17) | 0.397 |
|  | 2doses | **0.26 (0.06, 0.72)** | **0.025** | 0.54 (0.03, 3.01) | 0.568 |
| Sex | Female | Ref | Ref | Ref | Ref |
|  | Male | 1.05 (0.94, 1.18) | 0.402 | 1.00 (0.76, 1.34) | 0.979 |
| Age (years) Categorical | <1 | Ref | Ref | Ref | Ref |
|  | 1-<3 | **0.59 (0.50, 0.69)** | **<0.001** | 0.65 (0.42, 1.02) | 0.057 |
|  | 3+ | **0.49 (0.42, 0.57)** | **<0.001** | **0.44 (0.27, 0.71)** | **<0.001** |
| Prior Negative Tests | none | Ref | Ref | n/a | n/a |
|  | 1 to 3 | **0.70 (0.62, 0.79)** | **<0.001** | 0.98 (0.72, 1.33) | 0.877 |
|  | 4 or more | **0.64 (0.48, 0.85)** | **0.003** | 1.14 (0.68, 1.86) | 0.618 |
| Abbreviations: OR: odds ratio, CI: confidence interval, Ref: reference group  P-value<0.05 in **bold** | | | | | |

**Table S3.** Multivariable logistic regression model results, stratified by prior positivity, including age as continuous (model 1), in 5 categories (model 2), and as 3 categories (model 3), using most-recent post-vaccine eligible negative for controls.

| **Characteristic** | | **No Prior Positive Test (N = 5,959)** | | **Any Prior Positive Test (N = 1,206)** | |
| --- | --- | --- | --- | --- | --- |
| **Model 1** | | **OR (95% CI)** | **p-value** | **OR (95% CI)** | **p-value** |
| Vaccine Status | unvaccinated | Ref | Ref | Ref | Ref |
|  | 1dose | 0.79 (0.31, 1.73) | 0.577 | 0.38 (0.02, 2.04) | 0.364 |
|  | 2doses | **0.21 (0.05, 0.58)** | **0.009** | 0.49 (0.03, 2.64) | 0.496 |
| Sex | Female | Ref | Ref | Ref | Ref |
|  | Male | 1.05 (0.94, 1.18) | 0.376 | 1.00 (0.76, 1.33) | 0.986 |
| Age (years)  Continuous | One unit increase | **0.84 (0.81, 0.88)** | **<0.001** | **0.78 (0.70, 0.88)** | **<0.001** |
| Prior Negative Tests | none | Ref | Ref | Ref | Ref |
|  | 1 to 3 | **0.68 (0.60, 0.76)** | **<0.001** | 1.00 (0.73, 1.37) | 0.997 |
|  | 4 or more | **0.62 (0.47, 0.83)** | **0.001** | 1.19 (0.71, 1.96) | 0.495 |
| **Model 2** | | **OR (95% CI)** | **p-value** | **OR (95% CI)** | **p-value** |
| Vaccine Status | unvaccinated | Ref | Ref | Ref | Ref |
|  | 1dose | 0.81 (0.32, 1.79) | 0.632 | 0.41 (0.02, 2.17) | 0.393 |
|  | 2doses | **0.21 (0.05, 0.57)** | **0.009** | 0.51 (0.03, 2.75) | 0.522 |
| Sex | Female | Ref | Ref | Ref | Ref |
|  | Male | 1.05 (0.94, 1.18) | 0.395 | 1.00 (0.76, 1.33) | 0.977 |
| Age (years) Categorical | <1 | Ref | Ref | Ref | Ref |
|  | 1-<2 | **0.59 (0.50, 0.70)** | **<0.001** | 0.66 (0.41, 1.05) | 0.075 |
|  | 2-<3 | **0.52 (0.43, 0.63)** | **<0.001** | **0.51 (0.31, 0.86)** | **0.010** |
|  | 3-<4 | **0.46 (0.38, 0.55)** | **<0.001** | **0.44 (0.26, 0.75)** | **0.002** |
|  | 4+ | **0.47 (0.39, 0.57)** | **<0.001** | **0.34 (0.19, 0.59)** | **<0.001** |
| Prior Negative Tests | none | Ref | Ref | Ref | Ref |
|  | 1 to 3 | **0.70 (0.62, 0.79)** | **<0.001** | 1.01 (0.74, 1.38) | 0.959 |
|  | 4 or more | **0.66 (0.49, 0.88)** | **0.005** | 1.21 (0.72, 1.99) | 0.463 |
| **Model 3** | | **OR (95% CI)** | **p-value** | **OR (95% CI)** | **p-value** |
| Vaccine Status | unvaccinated | Ref | Ref | Ref | Ref |
|  | 1dose | 0.81 (0.32, 1.77) | 0.620 | 0.43 (0.02, 2.26) | 0.418 |
|  | 2doses | **0.21 (0.05, 0.57)** | **0.009** | 0.48 (0.03, 2.57) | 0.484 |
| Sex | Female | Ref | Ref | Ref | Ref |
|  | Male | 1.05 (0.94, 1.18) | 0.397 | 1.01 (0.76, 1.34) | 0.972 |
| Age (years) Categorical | <1 | Ref | Ref | Ref | Ref |
|  | 1-<3 | **0.56 (0.48, 0.65)** | **<0.001** | **0.60 (0.39, 0.94)** | **0.023** |
|  | 3+ | **0.47 (0.40, 0.55)** | **<0.001** | **0.39 (0.25, 0.64)** | **<0.001** |
| Prior Negative Tests | none | Ref | Ref | Ref | Ref |
|  | 1 to 3 | **0.70 (0.62, 0.79)** | **<0.001** | 0.99 (0.73, 1.36) | 0.962 |
|  | 4 or more | **0.65 (0.49, 0.86)** | **0.003** | 1.17 (0.70, 1.92) | 0.540 |

**Table S4.** Sensitivity analysis adding Health District to multivariable models, including age as continuous (model 1), in 5 categories (model 2), and as 3 categories (model 3).

| Characteristic | | Control is earliest eligible test | | Control is most-recent eligible test | |
| --- | --- | --- | --- | --- | --- |
| Model 1 | | **OR (95% CI)** | **p-value** | **OR (95% CI)** | **p-value** |
| Vaccine Status | unvaccinated | Ref | Ref | Ref | Ref |
|  | 1dose | 0.73 (0.31, 1.51) | 0.433 | 0.79 (0.34, 1.63) | 0.545 |
|  | 2doses | 0.36 (0.11, 0.910 | 0.055 | 0.28 (**0.09, 0.71)** | **0.017** |
| Sex | Female | Ref | Ref | Ref | Ref |
|  | Male | 1.05 (0.94, 1.17) | 0.359 | 1.05 (0.94, 1.17) | 0.361 |
| Age (years)  Continuous | One unit increase | **0.83 (0.80, 0.87)** | **<0.001** | **0.82 (0.79, 0.86)** | **<0.001** |
| Prior positives | none | Ref | Ref | Ref | Ref |
|  | 1 or more | **0.63 (0.54, 0.73)** | **<0.001** | **0.63 (0.54, 0.73)** | **<0.001** |
| Prior negative tests | none | Ref | Ref | Ref | Ref |
|  | 1 to 3 | **0.74 (0.66, 0.83)** | **<0.001** | **0.75 (0.67, 0.84)** | **<0.001** |
|  | 4 or more | **0.76 (0.59, 0.97)** | **0.031** | **0.77 (0.59, 0.98)** | **0.039** |
| Health District | Alleghany | Ref | Ref | Ref | Ref |
|  | Central Virginia | **3.83 (3.05, 4.82)** | **<0.001** | **3.86 (3.07, 4.86)** | **<0.001** |
|  | Cumberland Plateau | **2.46 (1.94, 3.13)** | **<0.001** | **2.50 (1.97, 3.17)** | **<0.001** |
|  | Lenowisco | 1.16 (0.92, 1.46) | 0.205 | 1.16 (0.93, 1.46) | 0.194 |
|  | Mount Rogers | **2.32 (1.89, 2.86)** | **<0.001** | **2.32 (1.89, 2.86)** | **<0.001** |
|  | New River | **1.45 (1.15, 1.83)** | **0.002** | **1.44 (1.15, 1.82)** | **0.002** |
|  | Pittsylvania/Danville | **1.50 (1.16, 1.93)** | **0.002** | **1.50 (1.16, 1.93)** | **0.002** |
|  | Roanoke City | **1.32 (1.00, 1.73)** | **0.047** | **1.33 (1.01, 1.74)** | **0.042** |
|  | West Piedmont | **1.91 (1.48, 2.46)** | **<0.001** | **1.91 (1.48, 2.46)** | **<0.001** |
| Characteristic | | **Control is earliest eligible test** | | **Control is most-recent eligible test** | |
| Model 2 | | **OR (95% CI)** | **p-value** | **OR (95% CI)** | **p-value** |
| Vaccine Status | unvaccinated | Ref | Ref | Ref | Ref |
|  | 1dose | 0.76 (0.32, 1.57) | 0.486 | 0.82 (0.35, 1.68) | 0.607 |
|  | 2doses | 0.35 (0.11, 0.89) | 0.050 | **0.28 (0.08, 0.70)** | **0.015** |
| Sex | Female | Ref | Ref | Ref | Ref |
|  | Male | 1.05 (0.94, 1.17) | 0.370 | 1.05 (0.94, 1.17) | 0.364 |
| Age (years)  Categorical | <1 | Ref | Ref | Ref | Ref |
|  | 1-<2 | **0.62 (0.53, 0.74)** | **<0.001** | **0.59 (0.50, 0.70)** | **<0.001** |
|  | 2-<3 | **0.51 (0.42, 0.60)** | **<0.001** | **0.49 (0.41, 0.58)** | **<0.001** |
|  | 3-<4 | **0.46 (0.38, 0.54)** | **<0.001** | **0.44 (0.37, 0.52)** | **<0.001** |
|  | 4+ | **0.46 (0.38, 0.55)** | **<0.001** | **0.43 (0.36, 0.51)** | **<0.001** |
| Prior positives | none | Ref | Ref | Ref | Ref |
|  | 1 or more | **0.64 (0.55, 0.75)** | **<0.001** | **0.64 (0.55, 0.75)** | **<0.001** |
| Prior negative tests | none | Ref | Ref | Ref | Ref |
|  | 1 to 3 | **0.77 (0.68, 0.86)** | **<0.001** | **0.77 (0.69, 0.87)** | **<0.001** |
|  | 4 or more | 0.80 (0.62, 1.02) | 0.080 | 0.81 (0.62, 1.04) | 0.097 |
| Health District | Alleghany | Ref | Ref | Ref | Ref |
|  | Central Virginia | **3.83 (3.05, 4.83)** | **<0.001** | **3.85 (3.06, 4.85)** | **<0.001** |
|  | Cumberland Plateau | **2.46 (1.93, 3.12)** | **<0.001** | **2.49 (1.96, 3.17)** | **<0.001** |
|  | Lenowisco | 1.15 (0.91, 1.44) | 0.240 | 1.15 (0.92, 1.45) | 0.225 |
|  | Mount Rogers | **2.32 (1.89, 2.85)** | **<0.001** | **2.31 (1.88, 2.85)** | **<0.001** |
|  | New River | **1.44 (1.15, 1.82)** | **0.002** | **1.44 (1.14, 1.82)** | **0.002** |
|  | Pittsylvania/Danville | **1.50 (1.17, 1.94)** | **0.002** | **1.51 (1.17, 1.94)** | **0.002** |
|  | Roanoke City | 1.31 (1.00, 1.72) | 0.052 | 1.32 (1.00, 1.73) | 0.050 |
|  | West Piedmont | **1.93 (1.49, 2.48)** | **<0.001** | **1.92 (1.49, 2.48)** | **<0.001** |
| Characteristic | | **Control is earliest eligible test** | | **Control is most-recent eligible test** | |
| Model 3 | | **OR (95% CI)** | **p-value** | **OR (95% CI)** | **p-value** |
| Vaccine Status | unvaccinated | Ref | Ref | Ref | Ref |
|  | 1dose | 0.76 (0.33, 1.57) | 0.489 | 0.82 (0.35, 1.70) | 0.618 |
|  | 2doses | **0.35 (0.11, 0.88)** | **0.048** | **0.28 (0.08, 0.69)** | **0.015** |
| Sex | Female | Ref | Ref | Ref | Ref |
|  | Male | 1.05 (0.94, 1.17) | 0.391 | 1.05 (0.94, 1.17) | 0.380 |
| Age (years)  Categorical | <1 | Ref | Ref | Ref | Ref |
|  | 1-<3 | **0.57 (0.49, 0.66)** | **<0.001** | **0.55 (0.47, 0.63)** | **<0.001** |
|  | 3+ | **0.46 (0.39, 0.53)** | **<0.001** | **0.44 (0.37, 0.51)** | **<0.001** |
| Prior positives | none | Ref | Ref | Ref | Ref |
|  | 1 or more | **0.65 (0.55, 0.75)** | **<0.001** | **0.65 (0.55, 0.76)** | **<0.001** |
| Prior negative tests | none | Ref | Ref | Ref | Ref |
|  | 1 to 3 | **0.76 (0.68, 0.86)** | **<0.001** | **0.77 (0.68, 0.86)** | **<0.001** |
|  | 4 or more | 0.78 (0.61, 1.00) | 0.056 | 0.79 (0.61, 1.02) | 0.070 |
| Health District | Alleghany | Ref | Ref | Ref | Ref |
|  | Central Virginia | **3.79 (3.02, 4.78)** | **<0.001** | **3.81 (3.03, 4.80)** | **<0.001** |
|  | Cumberland Plateau | **2.43 (1.91, 3.09)** | **<0.001** | **2.46 (1.94, 3.13)** | **<0.001** |
|  | Lenowisco | 1.14 (0.91, 1.44) | 0.246 | 1.15 (0.92, 1.45) | 0.232 |
|  | Mount Rogers | **2.31 (1.88, 2.85)** | **<0.001** | **2.30 (1.87, 2.84)** | **<0.001** |
|  | New River | **1.44 (1.14, 1.82)** | **0.002** | **1.44 (1.14, 1.82)** | **0.002** |
|  | Pittsylvania/Danville | **1.50 (1.16, 1.93)** | **0.002** | **1.50 (1.16, 1.93)** | **0.002** |
|  | Roanoke City | 1.31 (1.00, 1.73) | 0.051 | **1.32 (1.00, 1.73)** | **0.049** |
|  | West Piedmont | **1.92 (1.49, 2.47)** | **<0.001** | **1.91 (1.48, 2.47)** | **<0.001** |
| Abbreviations: OR: odds ratio, CI: confidence interval, Ref: reference group  P-value<0.05 in bold | | | | | |
